# Did Low Risk Perception Mediate the COVID-19 Second Wave in Bangladesh? A Cross-sectional Study on Risk Perception and Preventive Practice

**DOI:** 10.1101/2021.08.08.21257737

**Authors:** Farah Naz Rahman, AKM Fazlur Rahman, Shah Monir Hossain, Mohammad Abul Faiz, Abu Jamil Faisel, ATM Iqbal Anwar, Moudud Hossain, Tarek Mahmud Hussain, Tania Mahbub, Liaquat Ali

**Author notes:** **Correspondence to** Farah Naz Rahman, Office of the Executive Director, Centre for Injury Prevention and Research, Bangladesh, Address: B-162, Road-23 New DOHS, Mohakhali, Dhaka-1206, Bangladesh.

## Abstract

**Objective:** This study assessed the risk perception and preventive behavioral practice towards COVID-19 just prior to the second wave of corona, as well as the impact of perceived risk on preventive practices.

**Design, setting, participants, and outcome measures:** A cross-sectional study was conducted between December 2020 and January 2021, involving 1382 respondents aged 18 years and above from all eight divisions in Bangladesh. We used multiple linear regression to identify sociodemographic predictors of risk perception and multiple logistic regression to determine the relationship between risk perception and preventive practice.

**Results:** Low risk perception regarding COVID-19 was present among one-fifth of the respondents (19.8%). Younger age, being male, low education, single marital status, and rural residence were significantly associated with a low perceived risk of COVID-19. Hand washing and wearing mask were practiced by 80% and 67% of respondents, respectively. A low prevalence was noticed for social distancing (31%), avoiding social gathering (31%), and covering face while coughing/sneezing (18%). Furthermore, respondents with a high risk perception were found to be more likely than those with a low risk perception to practice all recommended COVID-19 preventive behaviors-hand washing (OR=2.4, 95% CI=1.5, 3.7), mask use (OR=3.4, 95% CI=2.3, 5), social distancing (OR=3.7, 95% CI=2.4, 5.6), sanitizer use (OR=2.7, 95% CI=1.8, 4.1), avoiding gathering (OR=2.3, 95% CI=1.6, 3.5), avoid touching face and mouth (OR=2.8, 95% CI=1.5, 5.3), and covering mouth while coughing/sneezing (OR=7, 95% CI=3.6, 13.4).

**Conclusion:** Considerable number of Bangladeshi adults had low risk perception and low practice of some vital COVID-19 preventive behaviors before the onset of second wave of corona. All preventive practices were also influenced by respondent’s risk perception. This highlights the importance of strengthening and optimizing risk communication strategy even when the number of corona cases are low.

**STRENGTHS AND LIMITATIONS OF THIS STUDY:**

- The study explored the perceived risk and preventive practices for COVID-19 in Bangladesh right before the recent onset COVID-19 second wave in the South Asian region, and included a larger sample size than previous studies.
- Unlike most other studies on COVID-19 risk perception that used online surveys, this study administered a face-to-face data collection from both urban and rural settings across all the eight divisions of Bangladesh.
- This is the first study in Bangladesh that investigated the effect of perceived risk of COVID-19 on the practice of a range of preventive behaviors, and used an analytical approach to quantify risk perception.
- Respondents’ self-reported information on COVID-19 preventive behavior practice is subject to be influenced by recall and desirability bias.
- The study was unable to explore the respondents’ frequency and adherence to preventive practices, as well as the influence of psychological factors on preventive behaviors.

## BACKGROUND

After a brief respite, the second wave of COVID-19 pandemic has begun to strike the densely populated Low and Middle Income Countries (LMIC) of South Asian region.[1–3] Bangladesh has also started experiencing the pandemic’s second wave, with the highest number of new cases (15,192) and deaths (247) recorded in a single day on 26^th^ July 2021.[4] Bangladesh detected its first COVID-19 case on 8^th^ March 2020. Following that, there was a spike in the number of corona cases from June to August 2020 (the first wave), which gradually declined until the first two months of 2021, with February recording the lowest single-day increase in cases in over nine months.[5] From this point forward, the number of daily positive corona cases has recently increased dramatically, marking the onset of second wave from the second week of March 2021. As a result, the country has implemented a strict lockdown from April 2021 to the present.

According to some national experts, people’s negligence in practicing preventive measures i.e. wearing mask, hand washing, social distancing, has pushed the country into this second wave of corona.[6] There is growing evidence that people’s perception of COVID-19 risk influences their use of preventive measures.[7–11] This brings up the ‘Peltzman hypothesis’, which, according to anecdotal reports, may have come in effect in South Asian countries; the theory implies that while protective measures are in place, people are more likely to be engaged in risky behaviors.[12,13] This theory may be applicable for Bangladesh as well, where the declining trend of corona cases, combined with the start of a vaccination drive, may cause a shift in risk perception and subsequent preventive practices, resulting in a resurgence of the second wave of infection. In accordance with this hypothesis, it is necessary to identify the pattern of perceived corona risks among the general population of Bangladesh just prior to the emergence of the second wave, in order to understand the possible driving forces behind this.

A couple of studies examined the risk perception regarding corona pandemic among the general population in Bangladesh. [14,15] However, these studies were conducted during the first wave of the pandemic and data were collected through online which limits the generalizability of the results to some extent. Furthermore, while there are several studies that investigated the relationship between risk perception and COVID-19 preventive practices, the majority of them are from the perspective of high-income countries.[7, 16–19] There is still little evidence regarding the influence of perceived risk of COVID-19 on the prevalence of preventive behavior of people in LMICs, particularly in the context of South Asian countries. This study investigated the prevalence and pattern of COVID-19 risk perception and preventive practices among the general population of Bangladesh around the onset of the pandemic’s second wave, as well as the impact of perceived risk on preventive behaviors, in order to fill a knowledge gap and facilitate risk communication strategy.

## METHODS

### Design, Setting, and Population

A cross-sectional survey was conducted among the population in selected urban and rural regions from eight divisions (major administrative units) of Bangladesh from December 2020 to January 2021. The divisions are-Dhaka, Chattogram, Mymensingh, Rajshahi, Khulna, Barishal, Sylhet, and Rangpur. The study population consisted of all adult Bangladeshi nationals aged 18 years and above from these regions.

### Sample Size and Sampling Technique

We randomly selected eight districts from each of the eight division: Dhaka, Coxs’ Bazaar, Patuakhali, Khulna, Sirajganj, Habiganj, Sherpur, and Rangpur. Two wards (elective unit of a city corporation) were randomly selected from each district headquarter or city corporation to collect data from urban dwellers. Subsequently, we randomly chose two villages from each district to collect data from rural respondents. Since urban Bangladesh have been holding more corona cases than the rural areas, we targeted 60 households from each selected ward and 45 households from each selected village. This resulted in a total target of 1680 households, which was greater than the required sample size at 80% power, 95% CI, 50% prevalence of high risk perception, design effect of 2, with gender stratification (male, female). The details of the sample size calculation and sampling procedure is provided in the Supplementary material.

### Data Collection and Ethical Considerations

Ten trained data collectors gathered data from the households. They chose the first household from the approximate geographical center of a ward or village. The following households were chosen in an anticlockwise direction. From each household an eligible respondent was randomly selected and approached for consent. We selected respondents who have been living in the selected ward or village for at least a year. Following informed written consent, the respondent was interviewed using a pretested semi-structured questionnaire. All data were collected with necessary COVID-19 safety precautions (e.g., personal protective equipment-gloves, mask, hand sanitizer, and social distancing). The institutional ethical review committee of Centre for Injury Prevention and Research Bangladesh (CIPRB) provided the ethical approval of this study [Ref: ERC/CIPRB/082020]. All ethical principles were followed throughout this research, as well as the guidelines for research during the pandemic issued by the Directorate General of Health Services Bangladesh (DGHS).

### Instrument

The instrument had three segments to obtain sociodemographic information, level of risk perception, and prevalence of preventive practices. The sociodemographic variables included age group (18-30, 31-45, 46-60, 60+ years), gender (male, female), education (no formal education, 1-5 years of schooling=primary, 6-10 years of schooling=secondary, >10 years of schooling=higher secondary and above), occupation (domestic work, service, minor business, medium and large business, agriculture, laborious work), marital status (currently married, never married/divorced/widow/widower), and location of residence (urban, rural).

We used seven statements with five Likert scale responses ranging from ‘strongly disagree’ to ‘strongly agree’ to explore the level of risk perception among the respondents. The statements were adapted from Dryhurst et al’s study on COVID-19 risk perception around the world and covered three risk dimensions: cognitive (likelihood), emotional (worry), and temporal-spatial dimensions [20]. A Cronbach’s Alpha coefficient of .84 validated the internal consistency of the scale. Furthermore, we asked respondents about their practice of 7 key COVID-19 related preventive behaviors in the last 7 days, which included hand washing, mask wearing, sanitizer use, social distancing, avoiding crowds, refraining from touching face/mouth, and covering mouth while coughing or sneezing. Respondents registered their self-reported practice and responded ‘Yes/No’ to these questions.

### Data Analysis

Of the 1680 households that were approached, 278 individuals (urban=159, rural=119) did not consent to participate and data from 20 respondents were excluded due to incomplete responses. This gave us a final sample of 1382 to be included in the analysis, resulting in a response rate of 82.3%.

We calculated a respondent’s risk perception score by assigning values to the five Likert scale responses (strongly disagree=1, disagree=2, Neutral=3, Agree=4, and Strongly Agree=5) and adding the results for seven statements, yielding an overall score ranging from 7 to 35. Table 1 lists the questions used to gather information on COVID-19 risk perception and preventive practices, along with their explanations.

**Table 1:** Survey Questions on Risk Perception and Preventive Practices Regarding COVID-19.

| Risk Perception of COVID-19 |  |  |
| --- | --- | --- |
| (Participants’ responses to risk statements was registered in a five-point Likert scale with score assigned for each response) |  |  |
| Category | Statements | Response & Assigned Score |
| Perceived Severity | COVID-19 is a severe disease | Strongly Agree=5<br>Agree=4<br>Neutral=3<br>Disagree=2<br>Strongly Disagree=1 |
|  | COVID-19 is harmful for my health |  |
| Perceived Susceptibility | If I do not adhere with the recommended health rules, I will get infected with COVID-19 |  |
|  | I have possibility to get infected with COVID-19 in the following 6 months |  |
| Emotion | I am tensed/worried about COVID-19 |  |
|  | I often think about COVID-19 |  |
|  | Information presented by media on COVID-19 is true/not overestimated |  |
| Preventive Behaviours for COVID-19 |  |  |
| (Respondents were asked if they practice the following activities in the last seven days) |  |  |
| Preventive Measure | Description | Response |
| Washing Hand | Handwashing with soap and water at least three times a day; before eating, after coming home from outdoors, after receiving any goods/packages that have arrived from outside your home, after going to the bathroom, and after coughing, sneezing, or blowing your nose. | Yes/No |
| Wearing Mask | Always putting on a mask while leaving the home/going to public places. | Yes/No |
| **Social Distancing** | Keeping a 3-feet distance from other people when interacting, with the exception of family members living in the same house. | Yes/No |
| **Using Hand Sanitizer** | Using alcohol-based hand sanitizer before eating food and after coughing/sneezing, in the absence of soap and water. | Yes/No |
| **Avoiding Social Gathering** | Refraining yourself from participating in any social gathering/close-contact settings and visiting any crowded place. | Yes/No |
| **Avoid Touching Face and Mouth** | Keeping your hands away from your eyes, nose, and mouth, particularly when you are outside home. | Yes/No |
| **Covering Mouth while Coughing/Sneezing** | While coughing/sneezing, covering your mouth by using a tissue or bending elbow. | Yes/No |

We converted the primary score to percentage by following a simple linear transformation [21]-

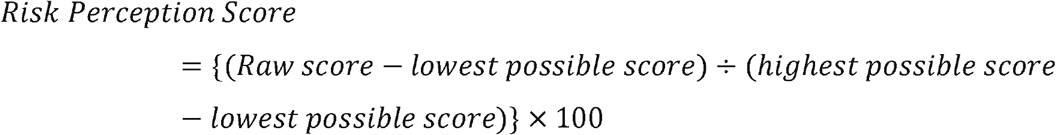

Following the equation, the new scores ranged from 0 to 100, where higher score indicated high risk perception and lower score indicated low risk perception. We then divided the score into three categories, with a score of up to 50 indicating low risk perception, a score of 51-75 indicating moderate risk perception, and a score of 76-100 indicating high risk perception. We conducted descriptive analysis to present the prevalence of level of risk perception and preventive practices among respondents. Subsequently, we used multivariate linear regression to identify the socio-demographic determinants of risk perception among respondents. In addition, we carried out multiple logistic regression to explore the relationship between risk perception and preventive practices. All of the preventive behaviors were considered as outcome variables, and separate regression analysis were conducted for each outcome. During analysis, we put sociodemographic variables along with risk perception as independent variables, however only reported the results for risk perception after adjusting for other covariates in the manuscript. All assumptions of regression analyses were met, including normal distribution of data, homogeneity of variance, and multicollinearity of variables.

### Patient and Public Involvement

After piloting of the initial questionnaire, it was modified with the consultation of the participants involved in pre-testing. We discussed and planned the research along with elected community leaders from the district headquarters of the selected districts. The findings have been shared with the Directorate General of Health Services (DGHS), Bangladesh, and will be widely disseminated through electronic media and journal articles.

## RESULTS

### Sociodemographic Characteristics of the Respondents

A total of 1382 individuals aged 18 years and above were surveyed in this study. Majority of the respondents were from younger and middle age group (73.8 % were less than or equal 45 years of age). Older adults (60+ years) constituted about 7% of the total respondents. Among the respondents, 51.5 % were males (ratio between males and females was 1.06:1) and majority (80.9%) were currently married. About one-third (34.1%) of respondents had a higher level of education (Higher secondary and above), while another 17.2% had no institutional education. The urban-to-rural ratio was 1.3: 1, where 57.4 % respondents were from urban areas. Nearly one-third (30.6 %) of respondents were engaged in agricultural or other laborious work, 21.7 % were engaged in minor business, and both service holders and domestic workers accounted for 16.1 % of total respondents. Detail percentages of all sociodemographic variables are provided in the supplementary material.

### Risk Perception of COVID-19 among Bangladeshi Adults

Participants recorded their responses for seven statements in a five Likert-scale (strongly agree/agree/neutral/disagree/strongly disagree). For better presentation, we grouped respondents who answered ‘strongly agree’ or ‘agree’ to a single category-‘Agreed,’ and similarly put respondents who answered ‘strongly disagree’ or ‘disagree’ to a single category-‘Disagreed’. These three levels of responses to the risk statements are depicted in Fig. 1. Percentages of five scale responses for all the seven statements are given in the supplementary material.

**Figure 1:**
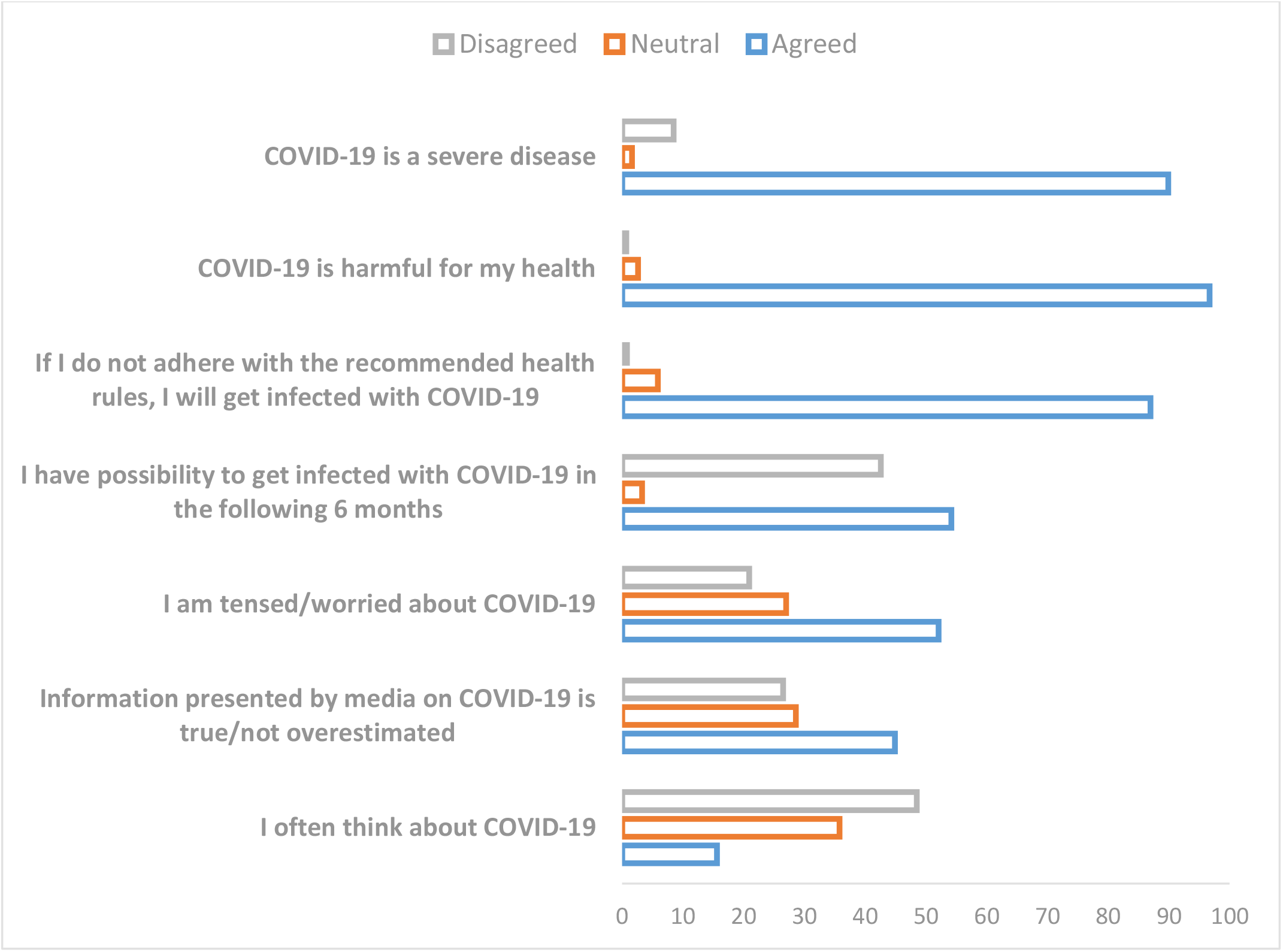
Responses of Bangladeshi Adults to Statements for Perceived COVID-19 Risk.(n=1382)

COVID-19 was perceived as a severe disease (89.9%) and as having a harmful effect (96.7%) on health by majority. More emphasis was seen on ‘harmfulness to individual health,’ with nearly two-thirds of respondents (64.5%) strongly agreeing with this statement (eTable-2). The practice of preventive behaviors was also regarded as important by majority (87%), as the respondents feel non-adherence to recommended health precautions can result in infection with COVID-19. Conversely, 42.6% respondents disagreed with the possibility of becoming infected with COVID-19 in the next 6 months, while a little more than half (54.2%) agreed. Although about half of the respondents (52.1%) agreed that they were worried about COVID-19, nearly half (48.5%) disagreed on having frequent thoughts about COVID-19 and its consequences. Regarding perception on COVID-19 information presented by media, about 45% respondents agreed with its authenticity, while 28.6% chose to be neutral.

A risk perception score was calculated (see methods), and the mean risk perception score of Bangladeshi adults was 67.1. Most of the respondents (46.5%) showed moderate level of risk perception which falls within the range of score 51-75. About 33% respondents were found to have a high level of risk perception (score 76-100), and the remaining one-fifth (19.8%) had low risk perception (score ≤50) regarding COVID-19. A Figure displaying the risk perception score of the respondents divided in three categories can be found in the in the supplementary material.

### Determinants of COVID-19 Risk Perception among Bangladeshi Adults

Mean risk perception scores of different sociodemographic groups along with the results of multiple linear regression analysis is presented in Table 2. This adjusted linear regression model predicts the effect of a socio-demographic factor on risk perception after controlling the effect of other covariates.

**Table 2.** The Association between Perceived COVID-19 Risk and Sociodemographic Factors among Bangladeshi Adults.

| **Adjusted Model** |  |  |  |  |
| --- | --- | --- | --- | --- |
| **Variables** | **Mean Risk Perception Score** | **B Coefficient** | **Sig** | **95% CI (Lower, Upper)** |
| **Age** |  |  |  |  |
| 18 to 30 years | 65.3 | Ref | Ref | Ref |
| 30 to 45 years | 65.7 | 4.25* | .00 | 1.98, 6.52 |
| 46 to 60 years | 69.6 | 9.90* | .00 | 7.10, 12.70 |
| 60+ years | 75.9 | 16.04* | .00 | 12.38, 19.70 |
| **Gender** |  |  |  |  |
| Female | 67.9 | Ref | Ref | Ref |
| Male | 66.3 | -3.68* | .00 | -5.51, -1.85 |
| **Marital Status** |  |  |  |  |
| Single/ Divorced / Widow | 70.7 | Ref | Ref | Ref |
| Currently Married/ Cohabiting | 66.2 | -3.35* | .00 | -5.83, -.86 |
| **Education** |  |  |  |  |
| No literacy | 58.7 | Ref | Ref | Ref |
| Primary | 65.1 | 7.96* | .00 | 5.12, 10.80 |
| Secondary | 66.5 | 10.48* | .00 | 7.58, 13.39 |
| Higher Secondary and above | 73.3 | 18.21* | .00 | 15.48, 20.94 |
| **Resident Location** |  |  |  |  |
| Rural | 71.9 | Ref | Ref | Ref |
| Urban | 63.5 | -9.59* | .00 | -11.41, -7.77 |

[Note: Adjusted model for multiple standardized linear regression coefficients. Variables adjusted are Age, Gender, Marital Status, Education, and Resident location. \*p< 0.01, N= 1382.]

Table-2 depicts that all sociodemographic variables-age, gender, education, marital status, and resident location have a significant association with risk perception score of Bangladeshi adults.

Increasing age resulted in higher perceived risk for COVID-19 among the respondents. The risk perception score of respondents aged 60+ years was 16.5 points higher than that of respondents aged 18-30 years. For respondents aged 30-45 years and 46-60 years, the risk perception score increased by 4.2 and 9.9 points respectively, when compared to 18-30 years’ age group. Similarly, a trend of higher perceived risk score with higher education was noticed. Compared to respondents with no formal education, respondents with higher secondary and above level education has 18.5 points higher risk perception score. Respondents having primary and secondary level education have also 7.9 and 10.4 points higher risk perception score respectively, than those with no formal education. Furthermore, males were found to have 3.6 points lower perceived risk score than females. Married respondents also had about 3 points lower risk perception than single/divorced/widow respondents. Moreover, compared to rural residents, the risk perception score among urban residents decreased by 9.59 points.

### Practice of Preventive Behaviors for COVID-19 among Bangladeshi Adults

Respondent’s self-reported practice of preventive behaviors related to COVID-19 is presented in Fig 2. The prevalence of practicing individual preventive behavior is shown for all respondents as well as for different risk perception levels.

**Figure 2:**
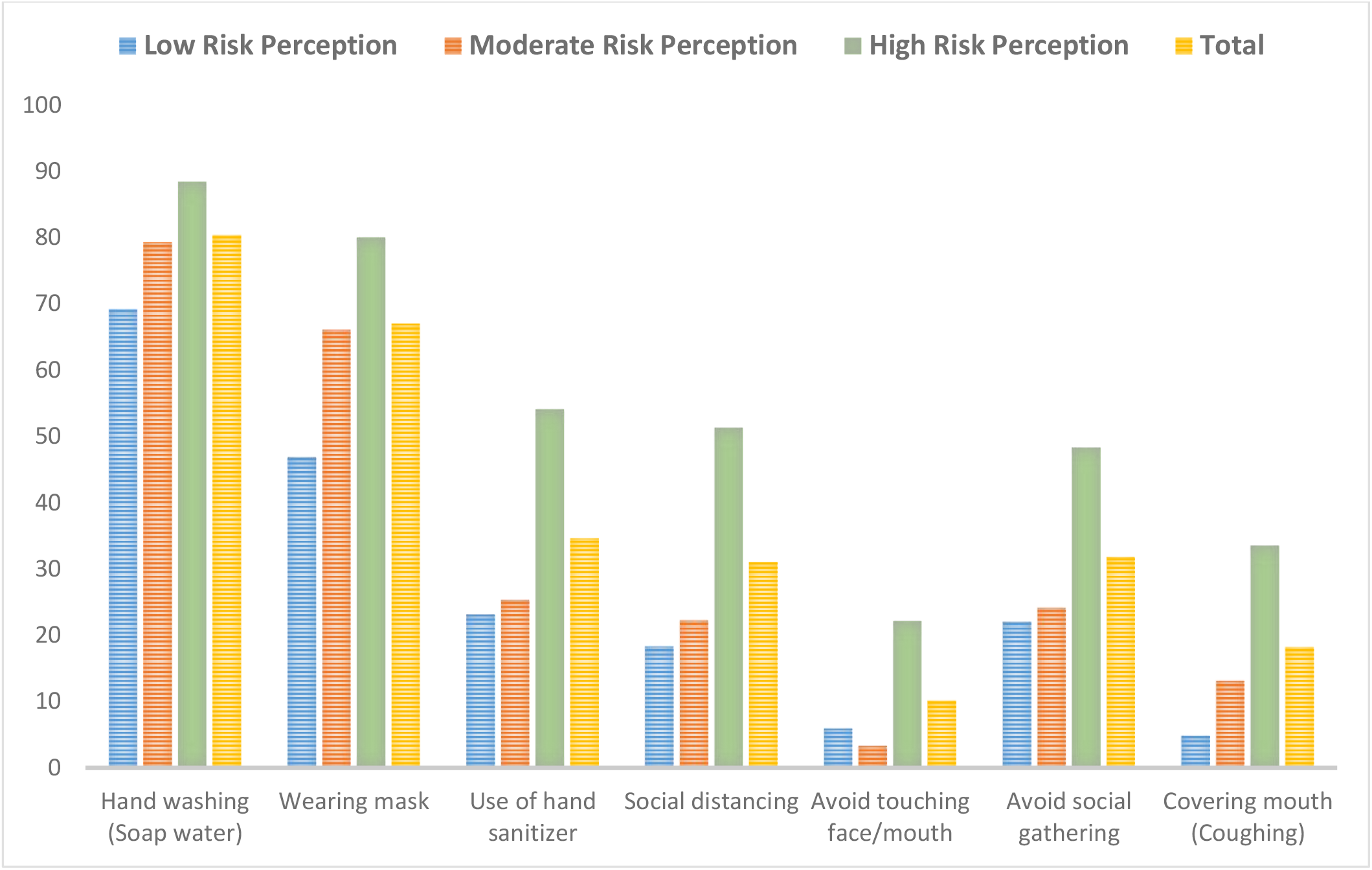
Prevalence of the Practice of Preventive Behaviors for COVID-19 among Bangladeshi Adults with Different Levels of Risk Perception.

The majority of respondents (80.4%) stated that they wash their hands with soap water for 20 seconds. Practice of using hand sanitizer, however, was present among only one-third (34.6%) of the respondents. Furthermore, two-thirds (67%) of the respondents reported of wearing mask while going outside. The practice of social distancing and avoiding social gatherings was comparatively low, with only about 31% of respondents admitting to engaging in these behaviors. The practice of covering one’s mouth while coughing or sneezing was also much lower (18.2%). Moreover, the practice of avoiding touching one’s face or mouth had the lowest prevalence (10.1%). For almost all preventive behaviors, the prevalence of practice was highest among respondents with high risk perception and lowest among respondents with low risk perception. The proportion of respondents engaged in social distancing, avoiding social gathering, and using sanitizer was more than double among the respondents with high risk perception than those in the other two risk perception categories.

### The Relationship between COVID-19 Risk Perception and Practice of Preventive Behaviors

Fig 3 summarizes the results of multiple logistic regression analyses predicting the effect of risk perception on preventive practices for COVID-19 after controlling the effect of sociodemographic variables-age, gender, education, occupation, marital status, and residence location. The figure depicts the calculated odds ratio (OR) of each behavioral outcome for moderate and high risk perception in comparison to low risk perception of COVID-19.

**Figure 3:**
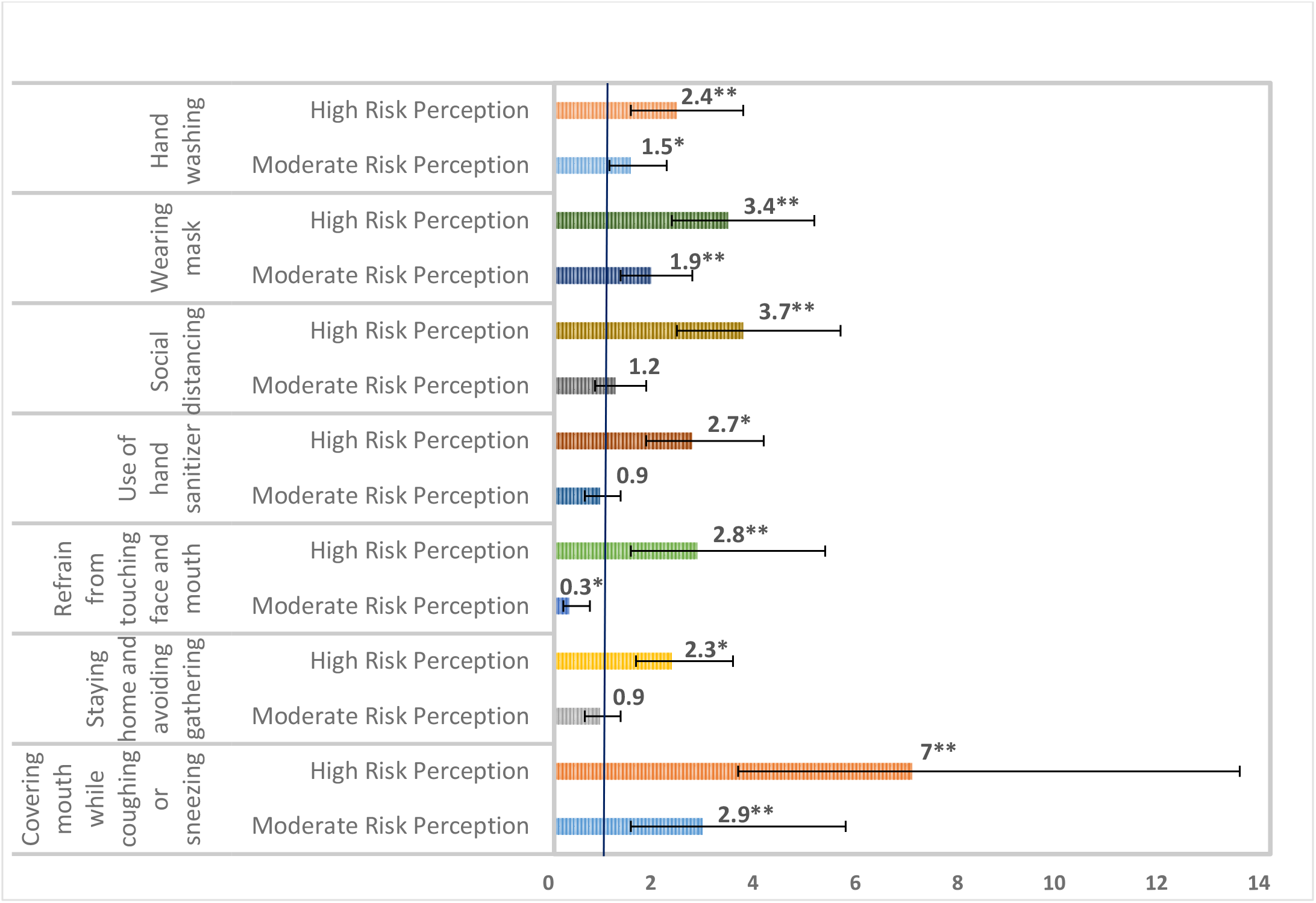
The Relationship between Perceived Risk of COVID-19 and Practice of Preventive Behaviors among Bangladeshi Adults. [Note: Adjusted OR from multiple logistic regression analyses are presented with error bars showing 95% CI. The vertical headings present preventive behavioral outcome (dependent variable) and the horizontal headings presents levels of risk perception (predictor variable). The odds of outcome were measured among respondents with high and moderate risk perception in comparison to respondents with low risk perception. Variables adjusted are age, gender, marital status, education, occupation, and residence location. *p<0.05, **p<0.01, N= 1382.]

When compared to respondents with a low risk perception, those with a high risk perception were 2.4 times more likely to wash their hands with soap water for 20 seconds, and those with a moderate risk perception were 1.5 times more likely to do so. Similarly, respondents with high and moderate risk perception respectively had 3.4 and 1.9 times higher odds of wearing mask, in comparison to those with low risk perception. Use of hand sanitizer, maintaining social distance, and avoidance of touching face/mouth, were also about three times more common among respondents with a high perceived risk than those with a low perceived risk. Furthermore, respondents with moderate to high risk perception were about three to seven times more likely than those with low risk perception to cover their mouth while coughing or sneezing. Moreover, tendency to avoid social gathering or crowded place were about two times higher among respondents with a high risk perception than those with a low risk perception. For better understanding, the results of multiple logistics regression analyses are also presented in tabulation form in the supplementary material.

## DISCUSSION

About one-fifth of the respondents (19.8%) had low risk perception regarding COVID-19, having a risk perception score of 50 or less on a scale of 0-100. Sociodemographic factors were significant predictors of risk perception, with older age, female gender, higher education, being married, and urban residence being associated with a higher perceived risk of COVID-19. Furthermore, all COVID-19 preventive behaviors such as hand washing, mask use, social distancing, sanitizer use, avoiding gathering, avoiding touching face/mouth, and covering mouth while coughing/sneezing, were found to be significantly associated with respondents’ level of risk perception.

Risk perception score was calculated based on respondents’ responses to seven statements. According to the responses to individual statements, the perceived severity of the disease was high, but the perceived susceptibility and emotional response to COVID-19 was much lower. Therefore, while majority of the respondents considered COVID-19 as a severe disease with harmful effect on health, only half were worried and believed they could become infected within the following 6 months. This is consistent with a COVID-19 risk survey in the Middle East reporting high perception for severity but low perception on susceptibility,[17] and with COVID-19 Knowledge, Attitude, and Practice (KAP) study in Bangladesh, in which 98.7% of respondents agreed that ‘COVID-19 is a dangerous disease.’[15] In contrast to our study, a higher prevalence of worriness were reported in a cross-sectional study in Bangladesh investigating COVID risk perception during the first wave, where 68.7% of respondents were very worried about COVID-19 and its consequences.[14] Another cross-sectional study conducted around the same time in Bangladesh used the Fear of COVID-19 scale and found that respondents had a moderate to high level of fear of contracting the disease.[22]

We found increasing age and higher education to be significantly associated with high perceived risk for COVID-19. Several studies in LMIC context also found a positive association between COVID-19 risk perception and participants’ age.[9,23]. Since older people are more likely to be affected by COVID-19 incidence and adverse outcomes, it stands to reason that their perceived risk is higher than that of other age groups. Our findings are further similar with Hossain et al.’s study in Bangladesh that found higher education to be associated with high fear score of contracting COVID-19.[22] As evidence suggests that knowledge of COVID-19 is positively associated with risk perception;[20,24] possible contribution of education to knowledge may resulted in a high risk perception. Additionally, we noticed a high perceived risk of COVID-19 among females, married participants, and rural residents. This is in line with the couple of studies in Bangladesh that found higher COVID-19 risk and fear among females.[14,22] Marital status wasn’t a significant predictor of perceived COVID-19 risk in Abir et al’s study in Bangladesh,[14] but it was found to be associated with high risk perception in a study in China.[24] Although few studies couldn’t establish a relation between geographical location and risk perception,[7,25] urban residents of our study revealed a significantly lower perceived risk than rural residents. It is concerning because the prevalence of COVID-19 is higher in urban areas in Bangladesh, necessitating special attention when developing risk communication programs.

A good practice regarding washing hand and wearing mask was noticed among the respondents, but the practice of maintaining social distance and avoiding social gathering was much lower. In general, we found that the practice of preventive behaviors was lower than previous studies conducted around the time of first wave of COVID-19 in Bangladesh. According to an online COVID-19 KAP survey in May 2020, the preventive practices of hand washing with soap and sanitizer, mask wearing, social distancing, and avoiding gathering were present among more than 90% of the respondents. [25] Along similar line, a couple of other studies exploring COVID-19 preventive practice reported high prevalence of mask use and social distancing in comparison to our study findings.[22,26] Moreover, we found that practice of preventive behaviors were more prevalent among respondents with higher perceived risk of COVID-19. This is supported by several other studies that demonstrated a positive relationship between risk perception and preventive practice.[9,11,16,18]

Our findings have few limitations. Due to data inadequacy, we couldn’t look at some important determinants of risk perception reported in other studies, such as presence of chronic condition and knowledge level of COVID-19. We couldn’t include wealth index in sociodemographic determinants as participants were found unwilling to respond to these questions in our piloting. Furthermore, we considered self-reported prevalence of preventive behaviors which is subject to recall and desirability bias. We were also unable to determine the frequency and adherence of these preventive behaviors among the respondents. Moreover, being a cross-sectional study, it couldn’t establish a causal relationship between COVID-19 risk perception and preventive practices. Despite these limitations, the study contributed to address a knowledge gap by exploring the relationship between COVID-19 risk perception and preventive practice in a vulnerable South Asian country context. It also used face-to-face data collection method from both rural and urban settings across all divisions of Bangladesh, and thus offers an improved generalizability compared to previous studies. This is possibly the first study to assess people’s perceived risk and practice just prior to the onset of COVID-19 second wave in South Asian region, and the findings conceptually support ‘Peltzmen Hypothsis’. Building on this evidence, future studies can use robust methodology to determine adherence to practice, its causal relationship with risk perception, and how it impacts the transmission of COVID-19 in community. In addition, exploratory researches can look in to the in-depth causes of reluctance of certain groups in practicing preventive measures for COVID-19.

As part of the National Preparedness and Response Plan (NPRP) for COVID-19 pandemic, some risk communication strategies have been developed and implemented in Bangladesh. However, these were mostly focused on disease severity and the health risk it carries. These programs’ influence may have contributed to the high perceived severity for COVID-19 found in our study and other similar studies. In contrast, we found that a considerable number of respondents had low risk perception, particularly in the domains of susceptibility and emotional response. Except hand washing and mask use, the prevalence of practicing all other recommended preventive health behavior was also present among less than one-third of the respondents. The prevalence of both perceived risk and preventive practices found in our study is comparatively lower than the previous studies conducted during the first wave of corona in Bangladesh. Additionally, we identified that a person’s level of risk perception has a significant impact on his practice of preventive behavior. Based on our findings and the experts’ proposed ‘Peltzman Effect,’ we hypothesize that the low incidence of corona cases after the first wave, confidence gained from surviving the first wave, and the onset of the vaccination drive, all together may have created a false sense of protection among Bangladeshi adults, leading them to engage in riskier behaviors. This is supported by the community mobility reports of google that showed an increased mobility in the recreational places such as shopping mall and restaurants, during the time of the study.[26] This was also higher than the mobility around the time of first wave of corona in Bangladesh. This indicates the need for optimizing and strengthening risk communication strategy of the country. These programs’ focus should be broadened to include a communication strategy for behavior change. This also highlights the importance of strategizing and continuing risk communication even when the COVID-19 incidences are low, to prevent future waves and similar catastrophes. Moreover, based on sociodemographic predictors, we identified community groups that are more likely to have a low perceived risk for COVID-19, which will allow the government to develop a targeted approach. We suggest incorporating community engagement, as well as the involvement of local elected representatives and community leaders, into the design and planning of targeted risk, social, and behavioral communication strategies.

## Data Availability

Data is publicly available.

https://figshare.com/articles/dataset/C19_Risk_perception_Preventive_Practice_sav/14661588

## ACKNOWLEDGEMENT

The authors would like to thank the CIPRB’s staff who helped in data collection.

## DATA SHARING STATEMENT

De-identified data is publicly available at <u>10.6084/m9.figshare.14661588</u>.

## ETHICS STATEMENT

The Institutional Ethical Review Committee (IRC) of Centre for Injury Prevention and Research Bangladesh (CIPRB) provided the ethical approval of this study [Ref: ERC/CIPRB/08052020].

## AUTHORS CONTRIBUTION

AKMFR, SMH, MAF, AJF, ATMIA, MH, TMH, and LA conceptualize the study. FNR and AKMF conducted the data analysis. FNR and TM prepared the first draft of the manuscript. The manuscript was finalized with the critical inputs of SMH, MAF, AJF, ATMIA, MH, TMH, and LA. All others reviewed and approved the final version.

