## supplementary material for "Did Low Risk Perception Mediate the COVID-19 Second Wave in Bangladesh? A Cross-sectional Study on Risk Perception and Preventive Practice"

**SAMPLE SIZE CALCULATION**

Sample size was calculated using the following formula:

$$n=\frac{Z^{2}P(1-P)}{d^{2}}$$

In absence of absolute data, we assumed that 50% of Bangladeshi adults will have high risk perception. The following parameters were used:

| · Prevalence of high risk perception among Bangladeshi adults (p) | 50% |
| --- | --- |
| · Confidence interval: | 95% |
| · Allowable error (d) | 0.05 |
| · Design effect: | 2 |
| · Response rate: | 95% |
| · Sex strata: | 2 |

Using these parameters, the preliminary sample size is 768. Further considering the sex strata, the number comes to 1536 (768*2) and for a 95% response rate, the required number that finally comes is 1612.

We used the following sampling technique to include the required number of individuals.

**Bangladesh**

Divisions (**n=8**)

Districts **(n=8**)

Villages [2 villages from each district, represents rural area]

Total= (8*2) = **16 villages**

Wards [2 wards from each district, represents urban area]

Total= (8*2) = **16 wards**

60 respondents from each ward

Total= (16*60) = **960 urban** respondents

45 respondents from each village

Total= (16*45) = **720 rural** respondents

Total Respondents approached=**1680**

**STEPS TO INCLUDE RESPONDENTS IN ANALYSIS**

**ENROLMENT**

**Total Number of Individuals Approached = 1680**

**Didn’t Provide Consent: 278**

**Total Number of Responses Included in Analysis:1382**

**Number of Respondents Participated in the Study: 1402**

**Number of Incomplete Responses: 20**

**SOCIODEMOGRAPHIC CHARACTERISTICS OF THE RESPONDENTS**

**Supplementary Table-1: Socio-demographic characteristics of the Bangladeshi Adults**

| **Characteristics of Respondents** | **Measurement of variables** | **N** | **Percentage (%) Total** |
| --- | --- | --- | --- |
| Age | Age 18 to 30 years | 449 | 32.5 |
|  | Age 31 to 45 years | 571 | 41.3 |
|  | Age 46 to 60 years | 255 | 18.5 |
|  | Age 60+ years | 107 | 7.7 |
| Gender | Male | 712 | 51.5 |
|  | Female | 670 | 48.5 |
| Marital Status | Currently Married | 242 | 80.9 |
|  | Single/Divorced/Widow | 158 | 19.1 |
| Residential Location | Urban | 792 | 57.4 |
|  | Rural | 590 | 42.6 |
| Education | No Literacy | 238 | 17.2 |
|  | Primary | 348 | 25.2 |
|  | Secondary | 325 | 23.5 |
|  | Higher Secondary and above | 471 | 34.1 |
| Occupation | Domestic work  Service  Minor business  Middle and large business  Agriculture  Laborious work  Others | 222  223  300  80  149  274  134 | 16.1  16.1  21.7  5.8  10.8  19.8  9.7 |

**RISK PERCEPTION AMONG BANGLADESHI ADULTS**

**Supplementary Table-2: Responses to Statements for Perceived Risk Regarding COVID-19 among Bangladeshi Adults.**

| Statement | Response | | | | |
| --- | --- | --- | --- | --- | --- |
|  | **Strongly Disagree**  **N (P)** | **Disagree**  **N (P)** | **Neutral**  **N (P)** | **Agree**  **N (P)** | **Strongly Agree**  **N (P)** |
| COVID-19 is a severe disease | 11 (.8) | 106 (7.7) | 22 (1.6) | 660 (47.8) | 583 (42.2) |
| COVID-19 is harmful for my health | 8 (0.6) | 1 (0.1) | 36 (2.6) | 445 (32.2) | 892 (64.5) |
| If I do not adhere with the recommended health rules, I will get infected with COVID-19 | 2 (0.1) | 9 (0.7) | 81 (5.9) | 885 (64) | 405 (29.3) |
| I have possibility to get infected with COVID-19 in the following 6 months | 113 (8.2) | 475 (34.4) | 45 (3.3) | 310 (22.4) | 439 (31.8) |
| I am tensed/worried about COVID-19 | 87 (6.3) | 202 (14.6) | 373 (27) | 427 (30.9) | 293 (21.2) |
| I often think about COVID-19 | 172 (12.4) | 499 (36.1) | 495 (35.8) | 198 (14.3) | 18 (1.3) |
| Information presented by media on COVID-19 is true/not overestimated | 93 (6.7) | 273 (19.8) | 395 (28.6) | 359 (26) | 262 (19) |

**LEVEL OF RISK PERCEPTION AMONG BANGLADESHI ADULTS**

**Supplementary Figure-1: Proportion of Different Levels of Risk Perception among Bangladeshi Adults**

|  |  |
| --- | --- |
| High Risk Perception | 33.7 |
| Moderate Risk Perception | 46.5 |
| Low Risk Perception | 19.8 |

**THE RELATIONSHIP BETWEEN COVID-19 RISK PERCEPTION AND PRACTICE OF PREVENTIVE BEHAVIORS**

Supplementary Table-3 presents the results of multiple logistic regression analyses predicting the effect of risk perception on preventive practices for COVID-19.

**Supplementary Table 3: The Relationship between Perceived Risk of COVID-19 and Practice of Preventive Behaviors among Bangladeshi Adults.**

| **Preventive Practices (Outcome variable)** | **Level of Risk Perception (Predictor variable)** | **B** | **Sig.** | **OR** | **95% CI (lower, upper)** |
| --- | --- | --- | --- | --- | --- |
| Hand washing (with soap water for 20 sec) | Moderate Risk Perception | .44 | .01 | **1.5*** | (1.08, 2.2) |
|  | High Risk Perception | .88 | .00 | **2.4**** | (1.5, 3.7) |
| Wearing mask | Moderate Risk Perception | .66 | .00 | **1.9**** | (1.3, 2.7) |
|  | High Risk Perception | 1.02 | .00 | **3.4**** | (2.3, 5.09) |
| Use of hand sanitizer | Moderate Risk Perception | -.06 | .73 | 0.9 | (.8, 1.8) |
|  | High Risk Perception | 1.01 | .00 | **2.7**** | (2.4, 5.6) |
| Social distancing | Moderate Risk Perception | .20 | .30 | 1.2 | (.6, 1.3) |
|  | High Risk Perception | 1.31 | .00 | **3.7**** | (1.8, 4.1) |
| Refrain from touching face and mouth | Moderate Risk Perception | -.99 | .01 | **0.3*** | (.1, .7) |
|  | High Risk Perception | 1.05 | .00 | **2.8**** | (1.5, 5.3) |
| Staying home and avoiding gathering | Moderate Risk Perception | -.06 | .72 | 0.9 | (0.6, 1.3) |
|  | High Risk Perception | .87 | .00 | **2.3**** | (1.6, 3.5) |
| Covering mouth while coughing or sneezing | Moderate Risk Perception | 1.09 | .00 | **2.9**** | (1.5, 5.7) |
|  | High Risk Perception | 1.95 | .00 | **7.04**** | (3.6, 13.4) |

*p<0.05, **p<0.01
